# The devil is in the details: Reporting and transparent research practices in sports medicine and orthopedic clinical trials

**DOI:** 10.1101/2021.07.20.21260565

**Authors:** Robert Schulz, Georg Langen, Robert Prill, Michael Cassel, Tracey Weissgerber

**Affiliations:** Berlin Institute of Health at Charité Universitätsmedizin, QUEST Center for Responsible Research, Berlin, Germany; University of Potsdam, Department of Sport and Health Sciences, Potsdam, Germany; Institute for Applied Training Science, Leipzig, Germany; Brandenburg Medical School Theodor Fontane, Department of Orthopedics and Traumatology, Brandenburg, Germany

## Abstract

**Introduction:** While transparent reporting of clinical trials is essential to assess the risk of bias and translate research findings into clinical practice, earlier studies have shown that deficiencies are common. This study examined current clinical trial reporting and transparent research practices in sports medicine and orthopedics.

**Methods:** The sample included clinical trials published in the top 25% of sports medicine and orthopedics journals over nine months. Two independent reviewers assessed pre-registration, open data and criteria related to scientific rigor, the study sample, and data analysis.

**Results:** The sample included 163 clinical trials from 27 journals. While the majority of trials mentioned rigor criteria, essential details were often missing. Sixty percent (confidence interval [CI] 53-68%) of trials reported sample size calculations, but only 32% (CI 25-39%) justified the expected effect size. Few trials indicated the blinding status of all main stakeholders (4%; CI 1-7%). Only 18% (CI 12-24%) included information on randomization type, method, and concealed allocation. Most trials reported participants’ sex/gender (95%; CI 92-98%) and information on inclusion and exclusion criteria (78%; CI 72-84%). Only 20% (CI 14-26%) of trials were pre-registered. No trials deposited data in open repositories.

**Conclusions:** These results will aid the sports medicine and orthopedics community in developing tailored interventions to improve reporting. While authors typically mention blinding, randomization and other factors, essential details are often missing. Greater acceptance of open science practices, like pre-registration and open data, is needed. These practices have been widely encouraged, we discuss systemic interventions that may improve clinical trial reporting.

**Registration:** https://doi.org/10.17605/OSF.IO/9648H

## Introduction

The overarching goal of medical research is to improve healthcare for patients, which requires the biomedical community to translate study outcomes into clinical practice (1). Clinical trials are central to this process, as properly conducted trials reduce the risk of bias and increase the likelihood that results about new treatments will be trustworthy, reproducible and generalizable (2,3). Clinical trials must be properly designed, conducted, and reported (4) to facilitate translation. Poorly designed and conducted trials may not be trustworthy or reproducible. This undermines public trust in biomedical research and raises concerns about whether the trial costs and patient risks were justified (5,6). Poor reporting makes it difficult to distinguish between trials with and without a high risk of bias.

To improve clinical trial reporting, the Consolidated Standards of Reporting Trials (CONSORT) guidelines (7,8) have been recommended by the ICMJE and widely disseminated by the EQUATOR network (9,10). While reporting has improved over time, major deficiencies that can impair translation are still common (11,12). Details needed to assess the risk of bias were missing from many published trials. More than half of all trials failed to address allocation concealment, and almost one third of studies did not address blinding of participants and personnel (12). Similarly, among randomized controlled trials published in the top five orthopedics journals, 60% failed to address the blinding status of the participants and 58% did not specify the number of participants included in the final analysis (13).

Orthopedics and sports medicine researchers have joined efforts to improve study design and reporting. Newly formed societies (14,15) and editorial series (16) focus on improving research quality in sports medicine and orthopedics. These efforts are urgently needed, as only 1% of studies in high-impact orthopedic journals reported all ten criteria needed for risk of bias assessment (13). In 42% of papers, risk of bias could not be assessed due to incomplete reporting (13). Incomplete reporting of exercise interventions (17) makes it impossible to implement interventions in clinical practice or to assess the appropriateness of the control intervention (18).

In sports medicine related fields, meta-researchers suggested that scientists may be using questionable research practices (Table 1) after observing overinflated effect sizes (19) and an unreasonably high number of papers that support the study hypothesis (20). Comprehensive reporting may prevent these biases or make them easier to detect. However, earlier studies have shown that reporting deficiencies are still common in orthopedics (13) and general medical journals (12,21).

**Table 1.** Terminology and concepts

|  |  |
| --- | --- |
| **Concept** |  |
| **Questionable research practices** | Questionable research practices are defined as “Design, analytic, or reporting practices that have been questioned because of the potential for the practice to be employed with the purpose of presenting biased evidence in favor of an assertion” (22) |
| **Selective reporting/ cherry picking** | The decision about whether to publish a study or parts of a study is based on the direction or statistical significance of the results (23,24). Pre-registration and Registered Reports may prevent selective reporting (25,26), which is also known as cherry picking. |
| **Publication bias** | The decision about whether to publish research findings depends on the strength and direction of the findings (27). The odds of publication are nearly four times higher among clinical trials with positive findings, compared to trials with negative or null findings (28). |
| **Outcome reporting bias** | Only particular outcome variables are included in publications and decisions about which variables to include are based on the statistical significance or direction of the results (23). Outcomes that are statistically significant have higher odds of being fully reported than non-significant outcomes (29,30). |
| **Attrition bias** | Attrition refers to reductions in the number of participants throughout the study due to withdrawals, dropouts, or protocol deviations. Attrition bias occurs when there are systematic differences between people who leave the study and those who continue (31).<br>For example, a trial shows no differences between two treatments. In one group, however, half the participants dropped out because they underwent surgery due to worsening symptoms. |
| **Null hypothesis statistical testing (NHST)** | NHST is originally based on theories of Fischer and Neyman-Pearson. The null hypothesis is rejected or accepted depending on the position of an observed value in a test distribution. While NHST is standard practice in many fields, the International Committee of Medical Journal Editors warns against the sole reliance on NHST due to several shortcomings of this approach (32). |
| **p-Hacking** | Describes the process of analyzing the data in multiple ways until statistically significant results are found. |
| **HARKing** | HARKing, or hypothesizing after results are known, is defined as presenting a post hoc hypothesis as if it were an a priori hypothesis (33). |

Therefore, this meta-research study examined reporting among clinical trials published in the top 25% of sports medicine and orthopedics journals. Our objective was to assess the prevalence of reporting for selected criteria, including pre-registration, open data and reporting of randomization, blinding, sample size calculations, data analysis and the flow of participants through the study. Meta-research data on clinical trial design, conduct and reporting will help researchers in sports medicine to implement targeted measures to improve trial design and reporting.

## Methods

### Protocol Pre-registration

The study was pre-registered on the Open Science Framework (RRID:SCR_003238) at https://doi.org/10.17605/OSF.IO/9648H. Additional details regarding sample selection and screening, data abstraction, and sample size calculation can be found in the supplemental materials.

### Sample selection and screening

We systematically examined clinical trials published in the top 25% of orthopedics and sports medicine journals over nine months. This sampling strategy provides a broad overview of practices in the field while including high-impact journals, which have the potential to drive change. Journals in the orthopedics and sports medicine category were selected based on the Scimago Journal Rank indicator (34) (supplementary methods). The top 25% of journals (n=65) were entered into the PubMed search with article type (clinical trial) and publication date (2019/12:2020/08) filters. The search was run on September 16, 2020. All articles (n=175 from 27 journals) were uploaded into Rayyan (RRID:SCR_017584; 35) for screening. Two reviewers (RS, GL) screened titles and abstracts to exclude articles that were obviously not clinical trials, as defined by the ICMJE. The ICMJE defines a clinical trial as any research project that “prospectively assigns people or a group of people to an intervention, with or without concurrent comparison or control groups, to study the relationship between a health-related intervention and a health outcome”(9). Two independent reviewers (RS, GL, RP) then performed full-text screening. All papers meeting the ICMJE clinical trial definition were included, whereas articles that did not meet the definition were excluded. Studies looking at both health-related and non-health-related outcomes were included but data abstraction focused on health-related outcomes only. Disagreements were resolved by consensus.

### Data abstraction

Two independent assessors (RS, GL, RP) reviewed each article and its supplemental files to evaluate the reporting of pre-specified criteria and extracted data using preformatted Excel spreadsheets. Table 2 presents the main criteria that were abstracted and a reason for their selection. The transparency and rigor criteria are based on CONSORT criteria for methods and results reporting (7,8). We also abstracted additional open science criteria, focusing on open access and open data (36,37). The abstraction protocol was deposited on the Open Science Framework (RRID:SCR_003238) at https://osf.io/q8b46/.

**Table 2.** Criteria for reporting and transparent research practices. The table shows specific questions used to assess each outcome criteria and provides a brief justification for why each criteria was selected.

| Category | Assessment | Rationale and Context |
| --- | --- | --- |
| **Sample Size calculation** | Was an a priori sample size calculation performed?<br>What type of sample size calculation was performed?<br>Did the authors provide a justification for the expected effect size? | - Low power is associated with high rates of spurious findings and overinflated effect sizes (38), and there is evidence for low median statistical power in rehabilitation research [40].<br>- A priori sample size calculations help to prevent underpowered trials, however, they are regularly performed inadequately. Common problems include failing to justify the expected treatment effect and not stating all values required for calculation (39). The majority of sample size calculations in rehabilitation trials are missing expected effect sizes (40). |
| **Randomization & concealed allocation** | Did the authors address whether randomization was used?<br>If so, were the randomization type and method mentioned?<br>Were the following details of the allocation concealment procedure addressed?<br>- Who generated the randomization sequence?<br>- Who enrolled participants?<br>- Who assigned participants to groups? | - Inadequate randomization and allocation concealment procedures introduce selection bias and are associated with increased odds of significant but spurious results (41) and overestimated treatment effects (42). |
| **Blinding** | Did the article include a statement on blinding?<br>Was the blinding status of each of the major stakeholders mentioned (participants, healthcare providers, outcome assessors, data analysts)?<br>Was each stakeholder group blinded? | - Blinding prevents ascertainment bias in clinical trials. A lack of blinding is associated with overinflated effect sizes (43). Terms like double-blind are ambiguous, interpreted differently, and don't provide reliable information on blinding of specific stakeholder groups (44). These terms should be abandoned in favor of reporting the blinding status of all relevant stakeholders (8). |
| **Flow of participants** | Were the inclusion and exclusion criteria clearly stated?<br>Did the authors define how many participants were excluded at each phase of the study and list reasons for exclusion?<br>Did the authors present this information in a flow chart? | - Detailed inclusion and exclusion criteria help the reader to assess generalizability.<br>- Knowing when and why participants dropped out or were removed from the study is essential to estimate attrition bias. |
| **Data analysis** | Was a study hypothesis presented and a primary outcome specified?<br>Was the hypothesis supported or rejected? | - Specifying the study hypothesis and the primary outcome prospectively safeguards against selective reporting. Discrepancies between the registration and the study report may indicate outcome switching, which favors statistically |
|  | If NHST was performed, were exact p-values, degrees of freedom, and the test statistics presented?<br>Were standardized effect sizes and their precision reported? | significant results and introduces selective reporting bias (45,46).<br>- Reporting the test statistic and degrees of freedom allows readers to identify misreported p-values. In 13% of psychology studies, meta-researchers detected mismatches between p-values and the reported test statistic and degrees of freedom that would affect statistical conclusions (47).<br>- Analyses should take the magnitude, confidence, and likelihood of an effect into account, instead of focusing on whether effects are statistically significant. Effect sizes show the magnitude of effects within a study, while standardized effect sizes allow for comparisons across studies (48). |
| **Data visualization** | Were bar graphs used to visualize continuous data? | - Using bar graphs to visualize continuous data is problematic because many different data distributions can lead to the same bar graph. The actual data may suggest different conclusions from the summary statistics alone (49,50). |
| **Intervention reporting** | What type of intervention was performed (e.g. exercise, physical therapy, surgery)?<br>For exercise interventions:<br>- Was monitoring of adherence to the intervention addressed?<br>- Were essential details needed to replicate the experimental and control interventions (e.g. frequency, intensity, volume, and type of exercise) provided? | - When clinical trials do not report details needed to implement the intervention, findings cannot be translated into clinical practice. The minority of exercise studies provided enough information to allow others to replicate interventions (51). The high prevalence of insufficient reporting led to the establishment of new intervention reporting guidelines (52,53).<br>- Adherence can effect intervention efficacy. Intervention effects can be up to three times larger in fully adherent participants compared to partly adherent participants (54). |
| **Transparency criteria** | Was the study registered or pre-registered?<br>Was a data availability statement included? Were the data publically available?<br>Was the study openly accessible? | - Half of researchers admit to selectively reporting results and presenting post hoc analyses as if they had been pre-specified (22). Pre-registration protects against this. Pre-registration (since 2005) and data availability statements (since 2018) are mandatory for clinical trials (55).<br>- Open access papers generate more media coverage and citations (56).<br>- Open data facilitates collaboration and benefits society (56). In 2017, 21% of 316 biomedical journals (57) and 28% of funders (58) required open data. |

### Protocol Deviations

For trials with exercise interventions, we assessed the frequency, intensity, and volume of exercise for experimental and control interventions. The protocol was modified if the control intervention did not involve exercise. Control interventions were rated as fully reported if the frequency, the content, and the duration was described. Control groups that received no intervention (e.g. wait-and-see) were rated as fully reported if the activity status or number of other treatments were monitored.

Trial registration statement assessments were amended to determine whether trials were registered prospectively or retrospectively. Two abstractors (RS, MP) assessed each trial registration. Trials were considered pre-registered if their registration was completed before the first participant was enrolled. Otherwise, the trial was classified as retrospectively registered. If the primary outcome was changed after the study began, the trial was classified as retrospectively registered.

### Statistical Analysis

This exploratory study assessed the prevalence of reporting for selected criteria in sports medicine and orthopedics clinical trials. Results are presented as the percentage of trials reporting each outcome measure, with a 95% confidence interval.

Odds ratios and their 95% confidence intervals were calculated to examine the relationship between the completeness of reporting and pre-registration, the use of flow charts, or the presence of sample size calculations and the completeness of reporting. Odds ratios were interpreted as unclear if the confidence interval included 1. These analyses were not pre-registered.

### Sample Size Calculation

This exploratory study does not require formal sample size calculations. However, we adhered to conventional sample size recommendations for exploratory designs and performed a precision-based sample size calculation to obtain rough estimates of relevant sample sizes (supplemental methods). Depending on different assumptions, a required sample size of 124 to 203 trials was estimated.

## Results

175 articles were screened, and 168 articles were reviewed from 27 sports medicine and orthopedics journals (Figure S1, Table S1). Eleven articles were excluded because they did not meet the ICMJE clinical trial criteria. One extended conference abstract was excluded because it was not a full-length research article. Analyses included the remaining 163 papers.

### Rigor and Sample Criteria

#### Sample size calculations

In trials not reporting a priori sample size calculation (Figure 1), 2% (CI 0-5%; n=4) reported that no sample size calculation was performed because the study was an exploratory pilot study. Among trials reporting sample size calculations (n=98), 53% (CI 43-63%; n=52) included a justification for the expected effect size. The remaining trials either presented no justification (39%; CI 23-42; n=32) or used arbitrary effect size thresholds (14%; CI 7-21%; n=14). Almost all sample size calculations were based on statistical power (93%; CI 88-98%; n=96). Two sample size calculations were based on precision (2%; CI 0-5%). No calculations were based on Bayes methods.

**Figure 1.**
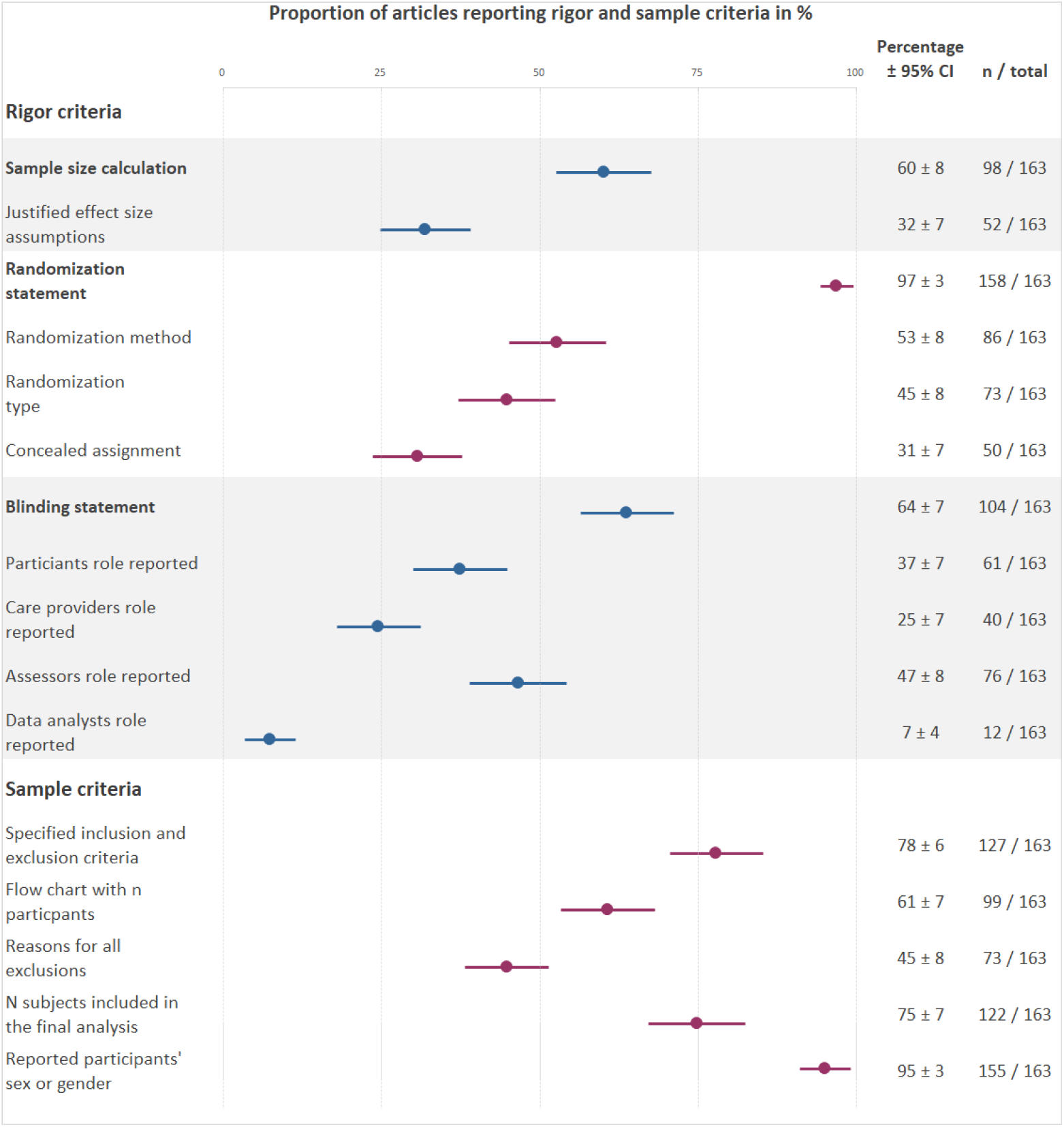
Reporting prevalence for rigor and sample criteria. This plot displays the percentage of trials that addressed each criteria. For information on the actual randomization or blinding status, please refer to the text. The different colored data points are for better visual differentiation of each subcategory.

#### Randomization and allocation concealment

In trials not addressing randomization (Figure 1), two trials (1%; CI 0-3%) were not randomized, and five trials did not mention randomization (3%; CI 0-6%).

Eight percent (CI 4-12%; n=13) of trials provided complete information on the allocation concealment procedure (defined as reporting who generated the randomization sequence, and who enrolled participants and assigned them to interventions). Some of this information was available 23% (CI 16-29%; n=37) of trials, and 69% (CI 62-76%; n=113) did not report any information. Few studies reported at least some information on all three factors needed to assess randomization and allocation concealment (randomization type, method, and allocation concealment; 18%; CI 12-24%; n=30).

#### Blinding

Two-thirds of trials addressed blinding (Figure 2). Among trials that addressed blinding (Figure 1), 81% (CI 73-88%; n=84) used blinding, while 19% (CI 12-27%; n=20) were not blinded. Only 4% (CI 1-7%; n=7) of all trials addressed the blinding status of all four stakeholder groups (Figure 2). Trials were most likely to address the blinding status of the outcome assessors and the participants. The blinding status of data analysts is typically unreported.

**Figure 2.**
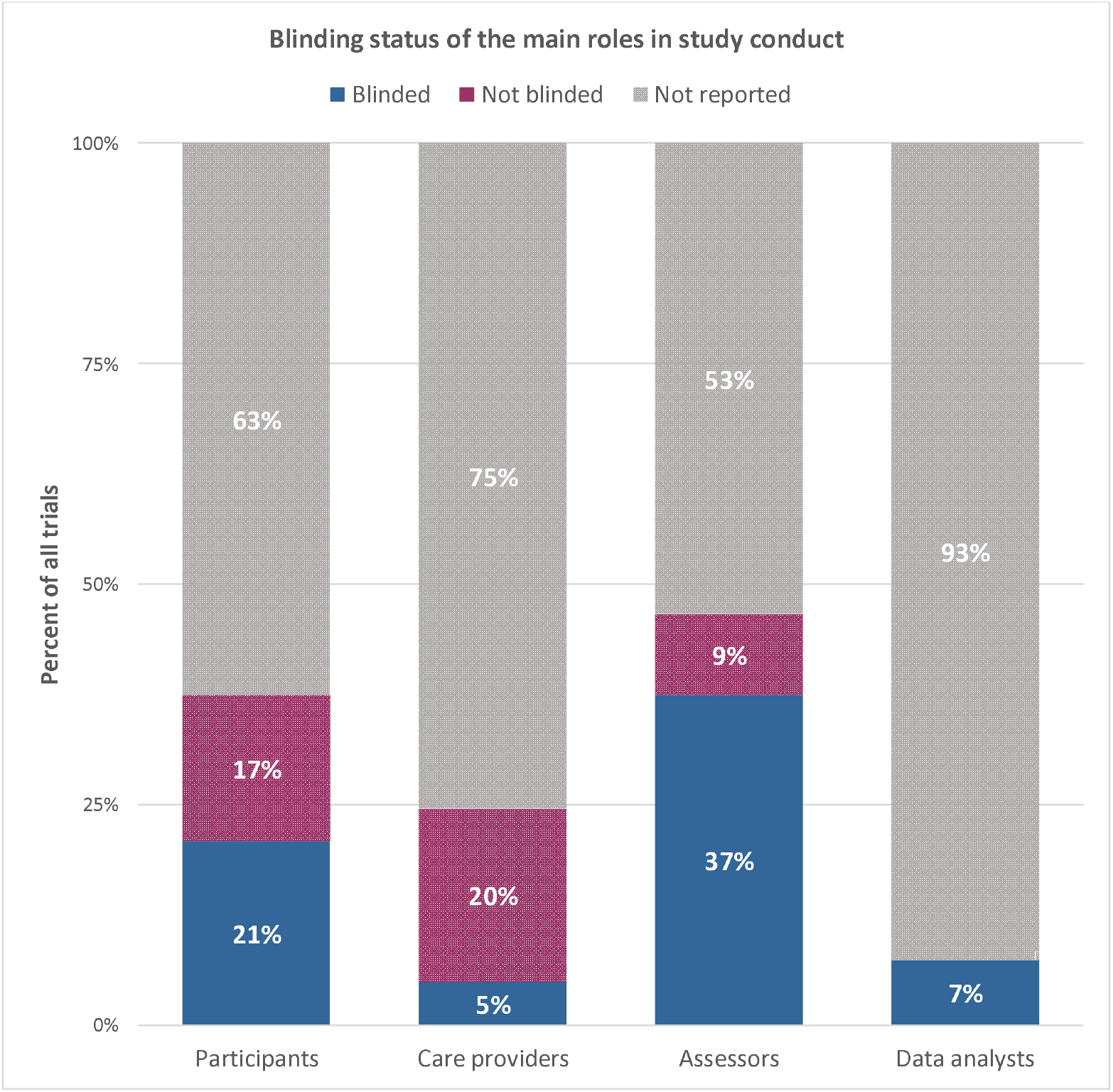
The blinding status across the main different stakeholder groups across all clinical trials (n=163)

### Sample-related Criteria

Approximately three-quarters of trials reported the inclusion and exclusion criteria and provided complete information on the number of participants at enrollment, after enrollment, and included in data analysis (Figure 1). Fewer trials used a flow chart to illustrate the number of included and excluded participants at each stage. Among trials that did not report the reasons for all exclusions after enrollment (Figure 1), 17% (CI 11-22%; n=24/90) reported the reasons for some exclusions and 33% (CI 26-41%; n=41/90) did not report any information.

In trials that stated participants’ sex or gender (Figure 1), a median of 51% (interquartile range (IQR) 27-71%) of participants were women in the group with the highest proportion of women, vs. 49% (IQR 22-66%) in the group with the lowest proportion of women.

### Intervention Criteria

The most frequent intervention type was exercise (44%; CI 37-52%; n=72), followed by surgery (26%; 19-32%; n=42). Diet (6%; CI 2-9%; n=9), physical therapy (5%; CI 2-8%; n=8), pharmacological interventions (4%;CI 0-2%; n=7) and manual therapy (1%; CI 0-2%; n=1) were uncommon. Fifteen percent (CI 9-20% n=24) of studies used other interventions.

We next examined reporting of details needed to assess or implement exercise interventions.Sixty-two percent (CI 50-73%; n=42) of trials with exercise interventions monitored adherence or compliance, one trial (1%; CI 0-4%) reported that adherence was not monitored, and 37% (CI 25-48%; n=25) of trials did not mention intervention adherence or compliance. All trials reported at least some information about the experimental exercise intervention, and most trials provided complete information (Table 2) (83%; CI 75-92%; n=60). Fewer trials reported complete information for the control interventions (63%; CI 51-74%; n=45). Five trials did not provide any information about the control intervention (7%; CI 1-13%).

### Data analysis and Transparency Criteria

#### Hypotheses and outcome measures

Nearly half of the articles specified a primary outcome and almost two-thirds of articles presented a hypothesis (Figure 3). Among clinical trials that reported a hypothesis (Figure 3), 61% (CI 53-68%; n=62) supported the main hypothesis, while 39% of trials (CI 32-47; n=40) did not support the main hypothesis.

**Figure 3.**
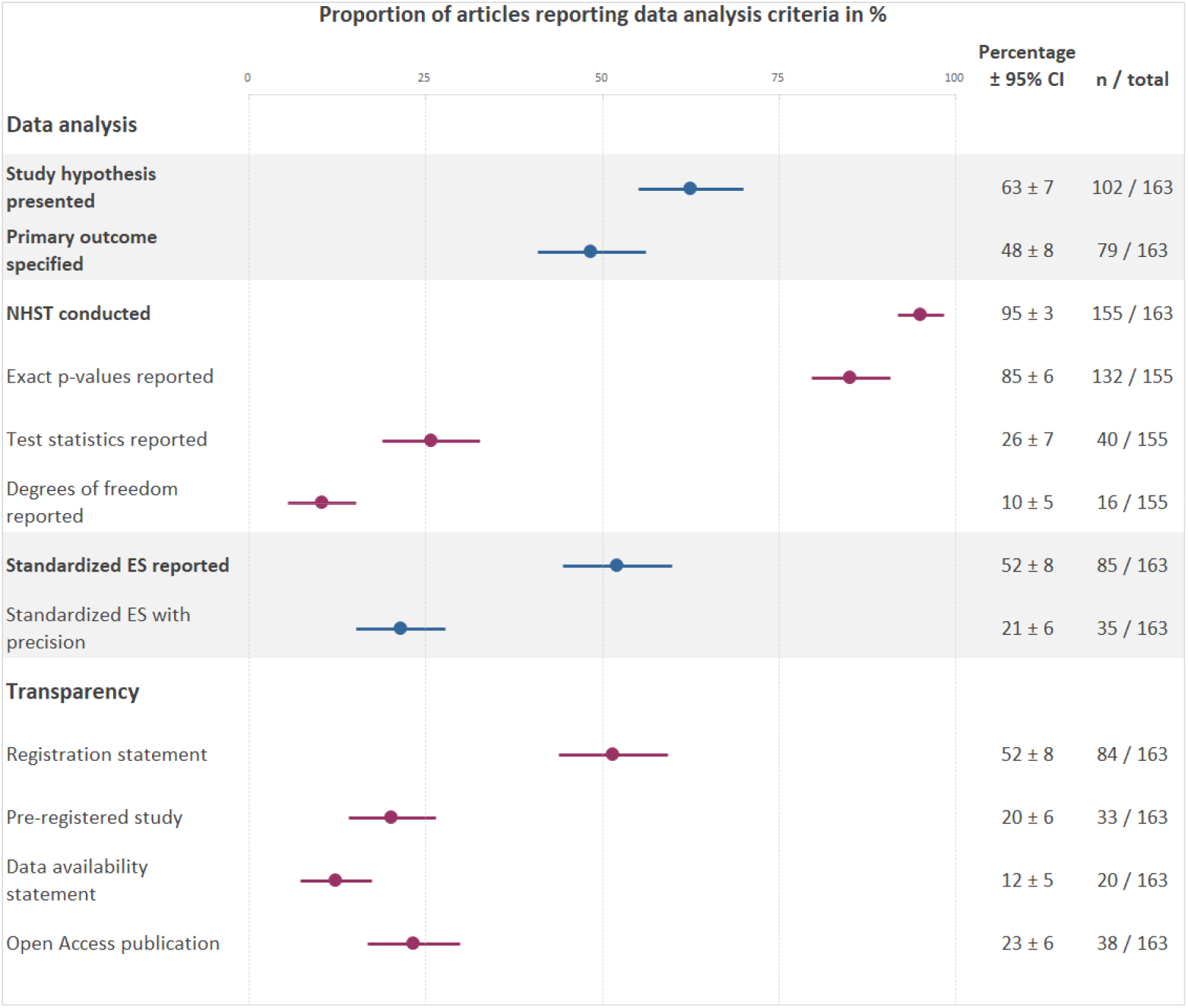
Reporting prevalence for data analysis and transparency criteria. This plot displays the percentage of trials that addressed each criteria. Abbreviations: NHST, null hypothesis statistical testing; ES, effect size

#### Statistical Reporting

Almost all studies used NHST (Figure 3). While most trials reported exact p-values, few reported test statistics and degrees of freedom. Approximately half of the trials reported standardized effect sizes but only 21% included the precision of the effect size estimates. One study reported Bayesian statistics (1%; CI 0-2%).

#### Data visualization

Bar graphs were used to display continuous data in 21% (CI 15-21%; n=34) of trials. These graphs should be replaced with more informative graphics (e.g. dot plots, box plots or violin plots) that show the data distribution(49,50).

### Transparency

Most of the studies with registration statements (Figure 3) were registered in ClinicalTrials.gov (n=52), followed by the Australian New Zealand Clinical Trials Registry (n=9), International Standard Randomized Controlled Trial Number Register (n=4), and other regional clinical trials registries (n=9). Less than half of the registered trials, and 20% of all trials, were pre-registered. The remaining trials with registration statements were registered retrospectively (58%; CI 48-69%; n=49/84). This included six prospectively registered trials where the primary outcome was changed after data collection started. Two studies with registration statements did not provide sufficient information to determine whether the study was registered prospectively or retrospectively (2%; CI 0-6%; n=2/84).

Data availability statements were uncommon (Figure 3). No trial with a data availability statement deposited data publically in an open repository. Twenty-one percent of trials with data availability statements (15-27%; n=4) noted that data were not publicly available, whereas 74% (67-80%; n=15) stated that data were available upon request. One study (5%; CI 2-9%) reported that all data were available in the main text and its supplements, however, raw data was not available in either location.

### Exploratory analyses

#### Pre-registration and reporting

Compared to unregistered or retrospectively registered studies, pre-registered studies were more likely to report complete information for randomization (type and method) and allocation concealment (OR 4.3; CI 1.9-10.0), whether all stakeholders were blinded (OR 8.6; CI 1.6-46.5), a priori sample size calculations (OR 2.5; CI 1.1-5.8), justifications for expected effect sizes used in power calculations (OR 2.5; CI 1.1-5.8), and specifying the primary outcome measure (OR 3.3; CI 1.5-7.1). The odds of reporting (OR 1.0; CI 0.48-2.1) or rejecting (OR 1.0; CI 0.42-2.6) the study hypothesis were not clearly different between unregistered and pre-registered studies.

#### Sample size calculations and reporting

The odds of rejecting the main hypothesis in trials with a priori sample calculations were unclear (OR 1.3; CI 0.6-2.8). Trials that provided justifications for the expected effect size were more likely to reject the study hypothesis (OR 2.5; CI 1.2-5.2).

#### Flow charts and reporting

The odds of reporting all reasons for dropouts (OR 4.6; CI 2.3-9.3) and explicitly reporting the number of participants in each group that were included in the data analysis (OR 163.3; CI 21.4-1248.5) were higher among studies that used flow charts to track participant flow, compared to those that did not.

## Discussion

Sports medicine and orthopedics researchers have recently emphasized rigorous study design and reporting to make research easier to understand, interpret, and translate into clinical practice (16). Calls for more transparent reporting in orthopedics and sports medicine (19,26,59) followed older studies suggesting that poor clinical trial reporting limits readers ability to assess study quality and risk of bias (13,60,61). Our study shows that while most studies include a general statement about rigor criteria, like blinding or randomization, these statements lack essential details needed to assess the risk of bias. The majority of trials report criteria related to the study sample, such as the sex of participants, inclusion and exlusion criteria, or the number of participants finally included in the analysis. Only 20% of studies were pre-registered. No study shared data in open repositories.

### Opportunities to improve reporting

These results highlight two main opportunities to improve transparency and reproducibility in sports medicine and orthopedics clinical trials; improving reporting for essential details of the main CONSORT elements and increasing uptake of open science practices.

First, our results indicate that most authors are aware that they need to address factors like blinding, randomization and sample size calculations; however, few provide the essential details required to evaluate the trial and interpret the results. Almost all trials addressed blinding, for example, but only 4% reported the blinding status of all main stakeholders. Educational efforts should emphasize the difference between informative and uninformative reporting (see example in Figure 4). CONSORT writing templates may also help (60). Target criteria should include the blinding status of all main stakeholders, randomization type and method, how and by whom concealed allocation was performed, and effect size justifications in sample size calculations.

**Figure 4.**
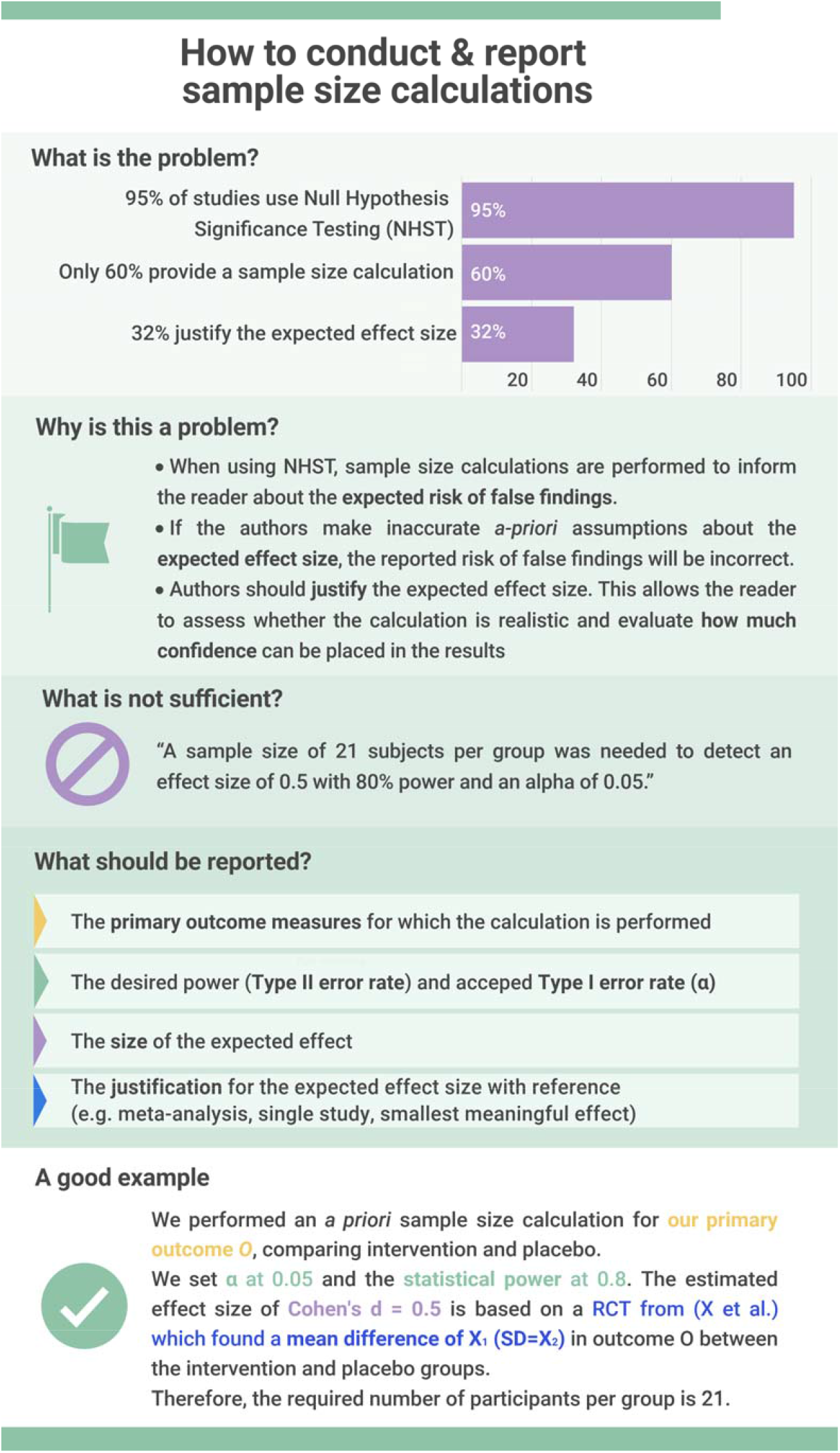
A priori sample size calculations are essential for generating meaningful results with clinical trials

Second, interventions are needed to increase pre-registration and data sharing. Although the ICJME has required clinical trial pre-registration since 2005 (61), only one-fifth of trials were pre-registered. Pre-registered studies had higher odds of reporting several rigor criteria, potentially suggesting that authors who preregister may be more aware of reporting guidelines. Our results are consistent with previous findings (62) that trial registrations were among the least reported CONSORT items in sports medicine. Sports medicine researchers have already noted that pre-registration and registered reports can prevent questionable research practice (26) (Table 1) or make them easier to detect (63).

Data were not shared in public repositories, suggesting that this topic requires special attention. The benefits of data sharing for authors include more citations (64,65), and increased opportunities to collaborate with researchers who want to perform secondary analyses (66). Recent materials have addressed many common concerns about sharing patient data, including data privacy and confidentiality (67–69). Regulations vary by country and institution. Some institutions have designated support staff for data sharing. Researchers should contact their institutions’ data privacy, statistics, or ethics offices to identify local experts. Seventy-four percent of trials with data availability statements noted that data were available on request. This is problematic, as such data are often unavailable and the odds of obtaining data decline precipitously with time since publication (70).

Interestingly, our exploratory analysis revealed that the odds of rejecting the study hypothesis were 2.5 (CI 1.2-5.2) times higher in trials that provided a justification for the expected effect size in sample size calculations. This might indicate overinflated effect sizes, as trials that based their sample size calculation on effect sizes published in earlier studies more often failed to find a similar sized effect. Inflated effect sizes were also observed in the psychological science reproducibility project, where replicated effects were generally smaller than those in the initial studies (71).

Authors should also be encouraged to report the data analysis transparently. Our study shows that reporting of test statistics and degrees of freedom yields much potential for improvement, as well as reporting of standardized effect sizes and their precision. Focusing on the magnitude and precision of differences, instead making decisions based on p-value thresholds, reduces the likelihood of spurious findings (72,73). Twenty-five to 38% of medical articles (74), and up to 50% in psychology papers (47), contain p-values that don’t match the reported test-statistic and degrees of freedom. These inaccurate p-values may alter study conclusions in 13% of psychology papers (47). Our study shows that these assessments are impossible in sports medicine and orthopedics clinical trials, as test statistics and degrees of freedom are rarely reported.

Reporting of criteria related to the study sample and to exercise interventions highlighted some positive points. Whereas Costello et al. (75) observed that less than 40% of sports and exercise study participants were females, indicating sex bias, our study, on average, shows an even distribution of sex/gender. Similarly the number of participants included in the analysis was reported in 75% of trials in the present study, compared to 42% of randomized controlled trials in orthopedic journals (13). The introduction of flow charts to display the participant flow in CONSORT 2010 may improve reporting for sample related criteria, as trials which included flow charts were more likely to report the number of participants included in the analysis and reasons for all exclusions. While the majority of studies reported key details of exercise interventions, reporting was less comprehensive for the control intervention and for intervention adherence or compliance.

### Options for systemic interventions to improve reporting

Ongoing reporting deficiencies in clinical trials highlight the need for systemic interventions to improve reporting. The 2010 CONSORT guideline has been endorsed by more than 50% of the core medical journals and the ICMJE (76). Persistent reporting deficiencies (12,21) indicate that endorsement without enforcement is insufficient (77,78), and engaging individuals, journals, funders, and institutions is necessary to improve reporting (79,80).

One option to improve reporting is for journals to enforce existing guidelines and policies. All journals in our sample were peer reviewed; yet there were major essential details were often missing from published trials. This suggests that peer review alone is insufficient. Alternatives include rigorous manual review by trained “trial reporting” assessors, automated screening or a combined approach. A journal program that trained early career researchers to check for common data visualization errors was well accepted by authors and increased compliance with data presentation guidelines (81). Implementing similar programs, using paid staff, could improve CONSORT compliance. Alternatively, automated screening tools may efficiently flag missing information for peer reviewers (82,83). Peer review systems at several journals include an automated tool that checks statistical reporting and guideline adherence (84). Tools are available to screen for risk of bias (RobotReviewer;RRID:SCR_021064 (85)), and CONSORT methodology criteria (CONSORT-TM;RRID:SCR_021051 (86)). The CONSORT tool performs well for frequently reported criteria, but needs more training data for less often reported criteria (86). New tools may be need to be created to assess details like the specifics of allocation concealment, blinding of specific stakeholders or justifications of expected effect sizes. As 52% of clinical trials in our sample were published in only five journals, systemic efforts to improve reporting on journal level can make a noteable difference on clinical trial reporting in the field.

A second option is automated screening of sports medicine and orthopedics preprints. Preprints, which are posted on public servers such as medRxiv and sportRxiv prior to peer review, allow authors to receive feedback and improve their manuscripts before journal submission. Large-scale automated screening of bioRxiv and medRxiv preprints for rigor and transparency criteria is feasible and could raise awareness about factors affecting transparency and reproducibility (87). Automated screening has limitations – the tools make mistakes and cannot always determine whether a particular item is relevant to a given study. Automated screening may complement peer review, but is not a replacement. The value of this approach will also depend on the proportion of trials that are posted as preprints.

Dashboards may offer a third option for improving reporting. Dashboards allow researchers to monitor changes over time and may incentivize transparent practices. Examples include dashboards on open science (88), and trial results reporting (89). In sports medicine and orthopedics, clinical trial dashboards could track transparent research practices for journals, society publishers, or all publications, and should include commonly missed items identified in this study. Researchers may need to develop new automated tools to track some criteria.

The scientific community has long relied on educational resources to improve reporting. On-demand resources include the CONSORT guideline use webinar by Altman (90), and open webinars on pre-registration, sample size justification and other topics offered by the Society for Transparency, Openness, and Replication in Kinesiology (91). Creating a single platform with field-relevant resources; then collaborating with large journals, publishers, and societies, may help to disseminate materials to the global orthopedics and sports medicine community.

### Limitations

Our CONSORT-based evaluation criteria for intervention reporting were not optimized for non-exercise or wait-and-see control interventions. While the assessments required by guidelines for intervention reporting (52,53) were beyond the scope of this study, previous studies assessed intervention reporting in detail (17,51,54,92). Larger, confirmatory studies are needed to examine relationships between different variables, as odds ratios calculated in the present study were exploratory post-hoc calculations. We examined the top 25% sports medicine and orthopedics journals; hence our findings may not be generalizable to journals that are not indexed by PubMed, lower tier journals, or non-English journals.

## Conclusions

Transparent reporting of clinical trials is essential to assess the risk of bias and translate research findings into clinical practice. Despite some improvements over time, older studies and studies in other fields show persisting deficiencies in clinical trial reporting. The present study in recent sports medicine and orthopedic clinical trials shows that authors often report general information on rigor criteria but few provide the essential details to assess risk of bias required by existing guidelines. Examples include the blinding status of all main stakeholders, information on the concealed assignment, or the justification of expected effect sizes in sample size calculations. Further, transparent research practies like pre-registration or data sharing are rarely used in sports medicine and orthopedics.

As reporting guidelines for clinical trial reporting are long established and well accepted across medical fields, the persisting lack of detailed reporting suggests that further interventions and different approaches are needed to improve clinical trial reporting further. We present different options for future interventions might investigate rigorous peer-reviewer training, automated screening of submitted manuscripts and preprints, and field-specific dashboards to moitor reporting and transparent research practies to increase awareness and track improvements over time. Our results show which aspects of clinical trial reporting have the greatest need for improvement. Researchers can use this data to tailor future interventions to improve reporting to the needs of the sports medicine and orthopedics community.

## Data Availability

All data are available on the OSF and may be accessed under the Creative Commons Attribution 4.0 International License at the following link: https://osf.io/q8b46/

https://doi.org/10.17605/OSF.IO/Q8B46

## Acknowledgements

We would like to acknowledge Mia Pattillo for her valuable contributions to extracting the registration status of trials.

## Competing Interests

All authors declare no competing interests.

## Notes

### Competing Interest Statement

The authors have declared no competing interest.

### Funding Statement

This study received no external funding.

### Author Declarations

Not applicable, because this article does not contain any studies with human or animal subjects.

### Summary of Updates

Author affiliations updated, typographical errors

## References

1. Emanuel EJ, Wendler D, Grady C. What makes clinical research ethical? JAMA. 2000;283(20):2701–11. doi:10.1001/jama.283.20.2701 Cited in: PubMed; PMID 10819955.

2. Califf RM, DeMets DL. Principles from clinical trials relevant to clinical practice: Part I. Circulation. 2002;106(8):1015–21. doi:10.1161/01.CIR.0000023260.78078.BB Cited in: PubMed; PMID 12186809.

3. Gerstein HC, McMurray J, Holman RR. Real-world studies no substitute for RCTs in establishing efficacy. The Lancet. 2019;393(10168):210–1. doi:10.1016/S0140-6736(18)32840-X

4. Zarin DA, Goodman SN, Kimmelman J. Harms From Uninformative Clinical Trials. JAMA. 2019. doi:10.1001/jama.2019.9892 Cited in: PubMed; PMID 31343666.

5. Feudtner C, Schreiner M, Lantos JD. Risks (and benefits) in comparative effectiveness research trials. N Engl J Med. 2013;369(10):892–4. doi:10.1056/NEJMp1309322 Cited in: PubMed; PMID 23964898.

6. van Delden JJM, van der Graaf R. Revised CIOMS International Ethical Guidelines for Health-Related Research Involving Humans. JAMA. 2017;317(2):135–6. doi:10.1001/jama.2016.18977 Cited in: PubMed; PMID 27923072.

7. Schulz KF, Altman DG, Moher D. CONSORT 2010 statement: updated guidelines for reporting parallel group randomised trials. PLoS Med. 2010;7(3):e1000251. doi:10.1371/journal.pmed.1000251 Cited in: PubMed; PMID 20352064.

8. Moher D, Hopewell S, Schulz KF, Montori V, Gøtzsche PC, Devereaux PJ, Elbourne D, Egger M, Altman DG. CONSORT 2010 explanation and elaboration: updated guidelines for reporting parallel group randomised trials. BMJ. 2010;340c869. doi:10.1136/bmj.c869 Cited in: PubMed; PMID 20332511.

9. ICMJE. Recommendations for the Conduct, Reporting, Editing and Publication of Scholarly Work in Medical Journals [Internet]. 2020 [cited 2020 Aug 4]. Available from: http://www.ICMJE.org.

10. Moher D, Simera I, Schulz KF, Hoey J, Altman DG. Helping editors, peer reviewers and authors improve the clarity, completeness and transparency of reporting health research. BMC Med. 2008;613. doi:10.1186/1741-7015-6-13 Cited in: PubMed; PMID 18558004.

11. Turner L, Shamseer L, Altman DG, Schulz KF, Moher D. Does use of the CONSORT Statement impact the completeness of reporting of randomised controlled trials published in medical journals? A Cochrane review. Syst Rev. 2012;160. doi:10.1186/2046-4053-1-60 Cited in: PubMed; PMID 23194585.

12. Dechartres A, Trinquart L, Atal I, Moher D, Dickersin K, Boutron I, Perrodeau E, Altman DG, Ravaud P. Evolution of poor reporting and inadequate methods over time in 20 920 randomised controlled trials included in Cochrane reviews: research on research study. BMJ. 2017;357j2490. doi:10.1136/bmj.j2490 Cited in: PubMed; PMID 28596181.

13. Chess LE, Gagnier J. Risk of bias of randomized controlled trials published in orthopaedic journals. BMC Med Res Methodol. 2013;1376. doi:10.1186/1471-2288-13-76 Cited in: PubMed; PMID 23758875.

14. Nielsen RO, Shrier I, Casals M, Nettel-Aguirre A, Møller M, Bolling C, Bittencourt NFN, Clarsen B, Wedderkopp N, Soligard T, Timpka T, Emery C, Bahr R, Jacobsson J, Whiteley R, Dahlstrom O, van Dyk N, Pluim BM, Stamatakis E, Palacios-Derflingher L, Fagerland MW, Khan KM, Ardern CL, Verhagen E. Statement on methods in sport injury research from the 1st METHODS MATTER Meeting, Copenhagen, 2019. Br J Sports Med. 2020;54(15):941. doi:10.1136/bjsports-2019-101323 Cited in: PubMed; PMID 32371524.

15. Zenko Z, Steele J, Mills J. Communications in Kinesiology: A new open access journal from the Society for Transparency, Openness, and Replication in Kinesiology; 2020. en.

16. Verhagen E, Stovitz SD, Mansournia MA, Nielsen RO, Shrier I. BJSM educational editorials: methods matter. Br J Sports Med. 2018;52(18):1159–60. doi:10.1136/bjsports-2017-097998 Cited in: PubMed; PMID 28818955.

17. Holden S, Rathleff MS, Jensen MB, Barton CJ. How can we implement exercise therapy for patellofemoral pain if we don’t know what was prescribed? A systematic review. Br J Sports Med. 2018;52(6):385. doi:10.1136/bjsports-2017-097547 Cited in: PubMed; PMID 29084726.

18. Losina E. Why past research successes do not translate to clinical reality: gaps in evidence on exercise program efficacy. Osteoarthritis Cartilage. 2019;27(1):1–2. doi:10.1016/j.joca.2018.09.006 Cited in: PubMed; PMID 30248501.

19. Knudson D. Confidence crisis of results in biomechanics research. Sports Biomech. 2017;16(4):425–33. doi:10.1080/14763141.2016.1246603 Cited in: PubMed; PMID 28632059.

20. Büttner F, Toomey E, McClean S, Roe M, Delahunt E. Are questionable research practices facilitating new discoveries in sport and exercise medicine? The proportion of supported hypotheses is implausibly high. Br J Sports Med. 2020. doi:10.1136/bjsports-2019-101863 Cited in: PubMed; PMID 32699001.

21. Kane RL, Wang J, Garrard J. Reporting in randomized clinical trials improved after adoption of the CONSORT statement. J Clin Epidemiol. 2007;60(3):241–9. doi:10.1016/j.jclinepi.2006.06.016 Cited in: PubMed; PMID 17292017.

22. Banks GC, O’Boyle EH, Pollack JM, White CD, Batchelor JH, Whelpley CE, Abston KA, Bennett AA, Adkins CL. Questions About Questionable Research Practices in the Field of Management. Journal of Management. 2016;42(1):5–20. doi:10.1177/0149206315619011

23. Hutton JL, Williamson PR. Bias in meta-analysis due to outcome variable selection within studies. Journal of the Royal Statistical Society: Series C (Applied Statistics). 2000;49(3):359–70. doi:10.1111/1467-9876.00197

24. Bernard R, Weissgerber TL, Bobrov E, Winham SJ, Dirnagl U, Riedel N. fiddle: a tool to combat publication bias by getting research out of the file drawer and into the scientific community. Clin Sci (Lond). 2020;134(20):2729–39. doi:10.1042/CS20201125 Cited in: PubMed; PMID 33111948.

25. Chambers C. What’s next for Registered Reports? Nature. 2019;573(7773):187–9. doi:10.1038/d41586-019-02674-6 Cited in: PubMed; PMID 31506624.

26. Caldwell AR, Vigotsky AD, Tenan MS, Radel R, Mellor DT, Kreutzer A, Lahart IM, Mills JP, Boisgontier MP. Moving Sport and Exercise Science Forward: A Call for the Adoption of More Transparent Research Practices. Sports Med. 2020;50(3):449–59. doi:10.1007/s40279-019-01227-1 Cited in: PubMed; PMID 32020542.

27. Chalmers l. Underreporting Research Is Scientific Misconduct. JAMA. 1990;263(10):1405. doi:10.1001/jama.1990.03440100121018

28. Hopewell S, Loudon K, Clarke MJ, Oxman AD, Dickersin K. Publication bias in clinical trials due to statistical significance or direction of trial results. Cochrane Database Syst Rev. 2009;(1):MR000006. doi:10.1002/14651858.MR000006.pub3 Cited in: PubMed; PMID 19160345.

29. Dwan K, Gamble C, Williamson PR, Kirkham JJ. Systematic review of the empirical evidence of study publication bias and outcome reporting bias - an updated review. PLoS ONE. 2013;8(7):e66844. doi:10.1371/journal.pone.0066844 Cited in: PubMed; PMID 23861749.

30. Kirkham JJ, Altman DG, Chan A-W, Gamble C, Dwan KM, Williamson PR. Outcome reporting bias in trials: a methodological approach for assessment and adjustment in systematic reviews. BMJ. 2018;362k3802. doi:10.1136/bmj.k3802 Cited in: PubMed; PMID 30266736.

31. Nunan D, Aronson J, Bankhead C. Catalogue of bias: attrition bias. BMJ Evid Based Med. 2018;23(1):21–2. doi:10.1136/ebmed-2017-110883 Cited in: PubMed; PMID 29367321.

32. ICMJE. Recommendations | Preparing a Manuscript for Submission to a Medical Journal: Methods - statistics [Internet]. 2021 [updated 2021 Apr 14; cited 2021 Apr 14]. Available from: http://www.icmje.org/recommendations/browse/manuscript-preparation/preparing-for-submission.html

33. Kerr NL. HARKing: hypothesizing after the results are known. Pers Soc Psychol Rev. 1998;2(3):196–217. doi:10.1207/s15327957pspr0203_4 Cited in: PubMed; PMID 15647155.

34. SCImago (nd). SJR — SCImago Journal & Country Rank [Portal] [Internet]. 2020 [cited 2021 Feb 22]. Available from: https://.scimagojr.com/journalrank.php?category=2732

35. Ouzzani M, Hammady H, Fedorowicz Z, Elmagarmid A. Rayyan-a web and mobile app for systematic reviews. Syst Rev. 2016;5(1):210. doi:10.1186/s13643-016-0384-4 Cited in: PubMed; PMID 27919275.

36. BIH QUEST. Open Science - BIH [Internet]. 2021 [updated 2021 Jan 22; cited 2021 Jan 22]. Available from: https://www.bihealth.org/en/research/quest-center/mission-approaches/open-science

37. Nosek BA, Alter G, Banks GC, Borsboom D, Bowman SD, Breckler SJ, Buck S, Chambers CD, Chin G, Christensen G, Contestabile M, Dafoe A, Eich E, Freese J, Glennerster R, Goroff D, Green DP, Hesse B, Humphreys M, Ishiyama J, Karlan D, Kraut A, Lupia A, Mabry P, Madon TA, Malhotra N, Mayo-Wilson E, McNutt M, Miguel E, Paluck EL, Simonsohn U, Soderberg C, Spellman BA, Turitto J, VandenBos G, Vazire S, Wagenmakers EJ, Wilson R, Yarkoni T. SCIENTIFIC STANDARDS. Promoting an open research culture. Science. 2015;348(6242):1422–5. doi:10.1126/science.aab2374 Cited in: PubMed; PMID 26113702.

38. Button KS, Ioannidis JPA, Mokrysz C, Nosek BA, Flint J, Robinson ESJ, Munafò MR. Power failure: why small sample size undermines the reliability of neuroscience. Nat Rev Neurosci. 2013;14(5):365–76. doi:10.1038/nrn3475 Cited in: PubMed; PMID 23571845.

39. Charles P, Giraudeau B, Dechartres A, Baron G, Ravaud P. Reporting of sample size calculation in randomised controlled trials: review. BMJ. 2009;338b1732. doi:10.1136/bmj.b1732 Cited in: PubMed; PMID 19435763.

40. Abdul Latif L, Daud Amadera JE, Pimentel D, Pimentel T, Fregni F. Sample size calculation in physical medicine and rehabilitation: a systematic review of reporting, characteristics, and results in randomized controlled trials. Arch Phys Med Rehabil. 2011;92(2):306–15. doi:10.1016/j.apmr.2010.10.003 Cited in: PubMed; PMID 21272730.

41. Hewitt C, Hahn S, Torgerson DJ, Watson J, Bland JM. Adequacy and reporting of allocation concealment: review of recent trials published in four general medical journals. BMJ. 2005;330(7499):1057–8. doi:10.1136/bmj.38413.576713.AE Cited in: PubMed; PMID 15760970.

42. Armijo-Olivo S, Saltaji H, da Costa BR, Fuentes J, Ha C, Cummings GG. What is the influence of randomisation sequence generation and allocation concealment on treatment effects of physical therapy trials? A meta-epidemiological study. BMJ Open. 2015;5(9):e008562. doi:10.1136/bmjopen-2015-008562 Cited in: PubMed; PMID 26338841.

43. Holman L, Head ML, Lanfear R, Jennions MD. Evidence of Experimental Bias in the Life Sciences: Why We Need Blind Data Recording. PLoS Biol. 2015;13(7):e1002190. doi:10.1371/journal.pbio.1002190 Cited in: PubMed; PMID 26154287.

44. Haahr MT, Hróbjartsson A. Who is blinded in randomized clinical trials? A study of 200 trials and a survey of authors. Clin Trials. 2006;3(4):360–5. doi:10.1177/1740774506069153 Cited in: PubMed; PMID 17060210.

45. Mathieu S, Boutron I, Moher D, Altman DG, Ravaud P. Comparison of registered and published primary outcomes in randomized controlled trials. JAMA. 2009;302(9):977–84. doi:10.1001/jama.2009.1242 Cited in: PubMed; PMID 19724045.

46. Chen T, Li C, Qin R, Wang Y, Yu D, Dodd J, Wang D, Cornelius V. Comparison of Clinical Trial Changes in Primary Outcome and Reported Intervention Effect Size Between Trial Registration and Publication. JAMA Netw Open. 2019;2(7):e197242. doi:10.1001/jamanetworkopen.2019.7242 Cited in: PubMed; PMID 31322690.

47. Nuijten MB, Hartgerink CHJ, van Assen MALM, Epskamp S, Wicherts JM. The prevalence of statistical reporting errors in psychology (1985-2013). Behav Res Methods. 2016;48(4):1205–26. doi:10.3758/s13428-015-0664-2 Cited in: PubMed; PMID 26497820.

48. Lakens D. Calculating and reporting effect sizes to facilitate cumulative science: a practical primer for t-tests and ANOVAs. Front Psychol. 2013;4863. doi:10.3389/fpsyg.2013.00863 Cited in: PubMed; PMID 24324449.

49. Weissgerber TL, Milic NM, Winham SJ, Garovic VD. Beyond bar and line graphs: time for a new data presentation paradigm. PLoS Biol. 2015;13(4):e1002128. doi:10.1371/journal.pbio.1002128 Cited in: PubMed; PMID 25901488.

50. Weissgerber TL, Winham SJ, Heinzen EP, Milin-Lazovic JS, Garcia-Valencia O, Bukumiric Z, Savic MD, Garovic VD, Milic NM. Reveal, Don’t Conceal: Transforming Data Visualization to Improve Transparency. Circulation. 2019;140(18):1506–18. doi:10.1161/CIRCULATIONAHA.118.037777 Cited in: PubMed; PMID 31657957.

51. Slade SC, Keating JL. Exercise prescription: a case for standardised reporting. Br J Sports Med. 2012;46(16):1110–3. doi:10.1136/bjsports-2011-090290 Cited in: PubMed; PMID 22089077.

52. Hoffmann TC, Glasziou PP, Boutron I, Milne R, Perera R, Moher D, Altman DG, Barbour V, Macdonald H, Johnston M, Lamb SE, Dixon-Woods M, McCulloch P, Wyatt JC, Chan A-W, Michie S. Better reporting of interventions: template for intervention description and replication (TIDieR) checklist and guide. BMJ. 2014;348g1687. doi:10.1136/bmj.g1687 Cited in: PubMed; PMID 24609605.

53. Slade SC, Dionne CE, Underwood M, Buchbinder R, Beck B, Bennell K, Brosseau L, Costa L, Cramp F, Cup E, Feehan L, Ferreira M, Forbes S, Glasziou P, Habets B, Harris S, Hay-Smith J, Hillier S, Hinman R, Holland A, Hondras M, Kelly G, Kent P, Lauret G-J, Long A, Maher C, Morso L, Osteras N, Peterson T, Quinlivan R, Rees K, Regnaux J-P, Rietberg M, Saunders D, Skoetz N, Sogaard K, Takken T, van Tulder M, Voet N, Ward L, White C. Consensus on Exercise Reporting Template (CERT): Modified Delphi Study. Phys Ther. 2016;96(10):1514–24. doi:10.2522/ptj.20150668 Cited in: PubMed; PMID 27149962.

54. Verhagen EALM, Hupperets MDW, Finch CF, van Mechelen W. The impact of adherence on sports injury prevention effect estimates in randomised controlled trials: looking beyond the CONSORT statement. J Sci Med Sport. 2011;14(4):287–92. doi:10.1016/j.jsams.2011.02.007 Cited in: PubMed; PMID 21429793.

55. ICMJE. Recommendations | Clinical Trials [Internet]. 2021 [updated 2021 Jan 19; cited 2021 Jan 19]. Available from: http://www.icmje.org/recommendations/browse/publishing-and-editorial-issues/clinical-trial-registration.html

56. McKiernan EC, Bourne PE, Brown CT, Buck S, Kenall A, Lin J, McDougall D, Nosek BA, Ram K, Soderberg CK, Spies JR, Thaney K, Updegrove A, Woo KH, Yarkoni T. How open science helps researchers succeed. Elife. 2016;5. doi:10.7554/eLife.16800 Cited in: PubMed; PMID 27387362.

57. Vasilevsky NA, Minnier J, Haendel MA, Champieux RE. Reproducible and reusable research: are journal data sharing policies meeting the mark? PeerJ. 2017;5e3208. doi:10.7717/peerj.3208 Cited in: PubMed; PMID 28462024.

58. European Comission. Facts and Figures for open research data: Figures and case studies related to accessing and reusing the data produced in the course of scientific production. [Internet]. 2019 [updated 2019 Nov 5; cited 2021 Apr 8]. Available from: https://ec.europa.eu/info/research-and-innovation/strategy/goals-research-and-innovation-policy/open-science/open-science-monitor/facts-and-figures-open-research-data_en#funderspolicies

59. Halperin I, Vigotsky AD, Foster C, Pyne DB. Strengthening the Practice of Exercise and Sport-Science Research. Int J Sports Physiol Perform. 2018;13(2):127–34. doi:10.1123/ijspp.2017-0322 Cited in: PubMed; PMID 28787228.

60. Barnes C, Boutron I, Giraudeau B, Porcher R, Altman DG, Ravaud P. Impact of an online writing aid tool for writing a randomized trial report: the COBWEB (Consort-based WEB tool) randomized controlled trial. BMC Med. 2015;13(1):221. doi:10.1186/s12916-015-0460-y Cited in: PubMed; PMID 26370288.

61. Angelis C de, Drazen JM, Frizelle FA, Haug C, Hoey J, Horton R, Kotzin S, Laine C, Marusic A, Overbeke AJPM, Schroeder TV, Sox HC, van der Weyden MB. Clinical trial registration: a statement from the International Committee of Medical Journal Editors. Ann Intern Med. 2004;141(6):477–8. doi:10.7326/0003-4819-141-6-200409210-00109 Cited in: PubMed; PMID 15355883.

62. Harris JD, Cvetanovich G, Erickson BJ, Abrams GD, Chahal J, Gupta AK, McCormick FM, Bach BR. Current status of evidence-based sports medicine. Arthroscopy. 2014;30(3):362–71. doi:10.1016/j.arthro.2013.11.015 Cited in: PubMed; PMID 24581261.

63. Warren M. First analysis of ‘pre-registered’ studies shows sharp rise in null findings. Nature. 2018. doi:10.1038/d41586-018-07118-1

64. Christensen G, Dafoe A, Miguel E, Moore DA, Rose AK. A study of the impact of data sharing on article citations using journal policies as a natural experiment. PLOS ONE. 2019;14(12):e0225883. doi:10.1371/journal.pone.0225883 Cited in: PubMed; PMID 31851689.

65. Colavizza G, Hrynaszkiewicz I, Staden I, Whitaker K, McGillivray B. The citation advantage of linking publications to research data. PLOS ONE. 2020;15(4):e0230416. doi:10.1371/journal.pone.0230416 Cited in: PubMed; PMID 32320428.

66. Lo B, DeMets DL. Incentives for Clinical Trialists to Share Data. N Engl J Med. 2016;375(12):1112–5. doi:10.1056/NEJMp1608351 Cited in: PubMed; PMID 27653562.

67. Mello MM, Francer JK, Wilenzick M, Teden P, Bierer BE, Barnes M. Preparing for responsible sharing of clinical trial data. N Engl J Med. 2013;369(17):1651–8. doi:10.1056/NEJMhle1309073 Cited in: PubMed; PMID 24144394.

68. Taichman DB, Sahni P, Pinborg A, Peiperl L, Laine C, James A, Hong S-T, Haileamlak A, Gollogly L, Godlee F, Frizelle FA, Florenzano F, Drazen JM, Bauchner H, Baethge C, Backus J. Data Sharing Statements for Clinical Trials - A Requirement of the International Committee of Medical Journal Editors. N Engl J Med. 2017;376(23):2277–9. doi:10.1056/NEJMe1705439 Cited in: PubMed; PMID 28581902.

69. Keerie C, Tuck C, Milne G, Eldridge S, Wright N, Lewis SC. Data sharing in clinical trials - practical guidance on anonymising trial datasets. Trials. 2018;19(1):25. doi:10.1186/s13063-017-2382-9 Cited in: PubMed; PMID 29321053.

70. Vines TH, Albert AYK, Andrew RL, Débarre F, Bock DG, Franklin MT, Gilbert KJ, Moore J-S, Renaut S, Rennison DJ. The availability of research data declines rapidly with article age. Curr Biol. 2014;24(1):94–7. doi:10.1016/j.cub.2013.11.014 Cited in: PubMed; PMID 24361065.

71. Nosek B. Estimating the reproducibility of psychological science. Science. 2015;349(6251):aac4716. doi:10.1126/science.aac4716 Cited in: PubMed; PMID 26315443.

72. Bernards JR, Sato K, Haff GG, Bazyler CD. Current Research and Statistical Practices in Sport Science and a Need for Change. Sports (Basel). 2017;5(4). doi:10.3390/sports5040087 Cited in: PubMed; PMID 29910447.

73. Riemann BL, Lininger MR. Principles of Statistics: What the Sports Medicine Professional Needs to Know. Clin Sports Med. 2018;37(3):375–86. doi:10.1016/j.csm.2018.03.004 Cited in: PubMed; PMID 29903380.

74. García-Berthou E, Alcaraz C. Incongruence between test statistics and P values in medical papers. BMC Med Res Methodol. 2004;4(1):13. doi:10.1186/1471-2288-4-13 Cited in: PubMed; PMID 15169550.

75. Costello JT, Bieuzen F, Bleakley CM. Where are all the female participants in Sports and Exercise Medicine research? Eur J Sport Sci. 2014;14(8):847–51. doi:10.1080/17461391.2014.911354 Cited in: PubMed; PMID 24766579.

76. CONSORT. Consort - Endorsers [Internet]. 2021 [updated 2021 Mar 19; cited 2021 Mar 19]. Available from: http://www.consort-statement.org/about-consort/endorsers1

77. Hirst A, Altman DG. Are peer reviewers encouraged to use reporting guidelines? A survey of 116 health research journals. PLoS ONE. 2012;7(4):e35621. doi:10.1371/journal.pone.0035621 Cited in: PubMed; PMID 22558178.

78. Shamseer L, Hopewell S, Altman DG, Moher D, Schulz KF. Update on the endorsement of CONSORT by high impact factor journals: a survey of journal “Instructions to Authors” in 2014. Trials. 2016;17(1):301. doi:10.1186/s13063-016-1408-z Cited in: PubMed; PMID 27343072.

79. MacLeod MR, Michie S, Roberts I, Dirnagl U, Chalmers I, Ioannidis JPA, Salman RA-S, Chan A-W, Glasziou P. Biomedical research: increasing value, reducing waste. The Lancet. 2014;383(9912):101–4. doi:10.1016/S0140-6736(13)62329-6

80. Moher D. Reporting guidelines: doing better for readers. BMC Med. 2018;16(1):233. doi:10.1186/s12916-018-1226-0 Cited in: PubMed; PMID 30545364.

81. Keehan KH, Gaffney MC, Zucker IH. CORP: Assessing author compliance with data presentation guidelines for manuscript figures. Am J Physiol Heart Circ Physiol. 2020;318(5):H1051–H1058. doi:10.1152/ajpheart.00071.2020 Cited in: PubMed; PMID 32196356.

82. Halffman W, Horbach SPJM. What are innovations in peer review and editorial assessment for? Genome Biol. 2020;21(1):87. doi:10.1186/s13059-020-02004-4 Cited in: PubMed; PMID 32362286.

83. Checco A, Bracciale L, Loreti P, Pinfield S, Bianchi G. AI-assisted peer review. Humanit Soc Sci Commun. 2021;8(1):1–11. En;en. doi:10.1057/s41599-020-00703-8

84. BMC. Advancing peer review at BMC [Internet]. 2021 [updated 2021 Apr 8; cited 2021 Apr 8]. Available from: https://www.biomedcentral.com/about/advancing-peer-review

85. Soboczenski F, Trikalinos TA, Kuiper J, Bias RG, Wallace BC, Marshall IJ. Machine learning to help researchers evaluate biases in clinical trials: a prospective, randomized user study. BMC Med Inform Decis Mak. 2019;19(1):96. doi:10.1186/s12911-019-0814-z Cited in: PubMed; PMID 31068178.

86. Kilicoglu H, Rosemblat G, Hoang L, Wadhwa S, Peng Z, Malički M, Schneider J, Ter Riet G. Toward assessing clinical trial publications for reporting transparency. J Biomed Inform. 2021;116103717. doi:10.1016/j.jbi.2021.103717 Cited in: PubMed; PMID 33647518.

87. Weissgerber T, Riedel N, Kilicoglu H, Labbé C, Eckmann P, Ter Riet G, Byrne J, Cabanac G, Capes-Davis A, Favier B, Saladi S, Grabitz P, Bannach-Brown A, Schulz R, McCann S, Bernard R, Bandrowski A. Automated screening of COVID-19 preprints: can we help authors to improve transparency and reproducibility? Nat Med. 2021;27(1):6–7. doi:10.1038/s41591-020-01203-7 Cited in: PubMed; PMID 33432174.

88. European Comission. Open science monitor [Internet]. 2018 [updated 2018 Nov 7; cited 2021 Apr 10]. Available from: https://ec.europa.eu/info/research-and-innovation/strategy/goals-research-and-innovation-policy/open-science/open-science-monitor_en

89. EU Trials Tracker. EU Trials Tracker — Who’s not sharing clinical trial results? [Internet]: Evidence-Based Medicine Data Lab; University of Oxford. 2021 [updated 2021 Apr 12; cited 2021 Apr 12]. Available from: https://eu.trialstracker.net/

90. Altman DG. WEBINAR: Doug Altman – CONSORT Statement guidance for reporting randomised trials | The EQUATOR Network [Internet]: EQUATOR. 2013 [updated 2021 Apr 8; cited 2021 Apr 8]. Available from: https://www.equator-network.org/2013/06/24/webinar-doug-altman-consort-statement-guidance-for-reporting-randomised-trials/

91. Society for Transparency, Openness, and Replication in Kinesiology. Stork - Resources [Internet]. 2021 [updated 2021 Apr 8; cited 2021 Apr 8]. Available from: https://storkinesiology.org/resources/

92. Hoffmann TC, Erueti C, Glasziou PP. Poor description of non-pharmacological interventions: analysis of consecutive sample of randomised trials. BMJ. 2013;347f3755. doi:10.1136/bmj.f3755 Cited in: PubMed; PMID 24021722.

