## Supplemental materials for "The devil is in the details: Reporting and transparent research practices in sports medicine and orthopedic clinical trials"

Supplemental material

**Methods**

**Sample Size Calculation**

This exploratory study does not require formal sample size calculations. However, conventionally, sample sizes between 30 and 150 subjects or items are recommended for exploratory study designs with non-probability sampling (1).

For information purposes only, a precision-based sample size estimation was performed to obtain rough estimates of relevant sample sizes. We assumed that three-quarters of articles would report the criteria (0.75), the margin of error would be 0.05, and a level of confidence of 0.8. These assumptions result in a calculated sample size of 124 articles. The estimated proportion was based on previous investigations in general medical journals (2–5). While the reporting prevalence varied substantially depending on the criterion, we chose an estimated reporting proportion of 75%, as the proportion of trials reporting information for risk of bias assessment was between 60 and 80% for most rigor criteria, and the latest large analysis of reporting in RCTs suggested that reporting was improving over time (5).

As the values chosen were estimates, additional sample size calculations were performed by varying the basic assumptions. The first alternative was to reduce the expected proportion from 75% to 66% (resulting in n=148) or 50% (resulting in n=165). Increasing the level of confidence from 0.8 to 0.9, with an expected proportion of 75%, would require an n of 203. After reviewing these estimates, the target sample size was set at approximately n=175 clinical trials. Sample size calculations were performed with the web-based application Statulator (RRID:SCR_021003; 6).

We searched for clinical trials published in August 2020; then went backward in time adding additional months until the target sample size was reached. The final search dates included clinical trials published between January and August 2020.

**Sample selection and screening process**

Journals were selected on basis of the Scimago journal ranking list from 2019 in the subject category orthopedics and sports medicine as determined by 2019 by Scimago Journal Rank indicator (7). The Scimago journal-ranking list was sorted by the Scientific Journal Ranking. The top 25% of journals (n=65) were then entered into the PubMed search with filters for article type (clinical trial) and publication date (2019/12:2020/08). The search was run on September 16, 2020.

The search string was:

(((((((((((((((((((((((((((((((((((((((((((((((((((((((((((((((("British journal of sports medicine"[Journal]) OR ("Sports Med"[jour])) OR ("The American journal of sports medicine"[Journal])) OR ("The bone & joint journal"[Journal])) OR ("The Journal of arthroplasty"[Journal])) OR ("The Journal of bone and joint surgery. American volume"[Journal])) OR ("Arthroscopy : the journal of arthroscopic & related surgery : official publication of the Arthroscopy Association of North America and the International Arthroscopy Association"[Journal])) OR ("Journal of bone and mineral research : the official journal of the American Society for Bone and Mineral Research"[Journal])) OR ("J Cachexia Sarcopenia Muscle"[jour])) OR ("Journal of shoulder and elbow surgery"[Journal])) OR ("Medicine and science in sports and exercise"[Journal])) OR ("Osteoarthritis and cartilage"[Journal])) OR ("International journal of sports physiology and performance"[Journal])) OR ("Knee surgery, sports traumatology, arthroscopy : official journal of the ESSKA"[Journal])) OR ("Skeletal muscle"[Journal])) OR ("Exercise and sport sciences reviews"[Journal])) OR ("Acta orthopaedica"[Journal])) OR ("Spine"[Journal])) OR ("International orthopaedics"[Journal])) OR ("Clinical orthopaedics and related research"[Journal])) OR ("Foot & ankle international"[Journal])) OR ("Therapeutic advances in musculoskeletal disease"[Journal])) OR ("Journal of science and medicine in sport"[Journal])) OR ("Orthopaedic journal of sports medicine"[Journal])) OR ("European spine journal : official publication of the European Spine Society, the European Spinal Deformity Society, and the European Section of the Cervical Spine Research Society"[Journal])) OR ("Scandinavian journal of medicine & science in sports"[Journal])) OR ("Bone & joint research"[Journal])) OR ("Current reviews in musculoskeletal medicine"[Journal])) OR ("Global spine journal"[Journal])) OR ("The Journal of the American Academy of Orthopaedic Surgeons"[Journal])) OR ("The Journal of hand surgery"[Journal])) OR ("Journal of teaching in physical education : JTPE"[Journal])) OR ("International journal of sport nutrition and exercise metabolism"[Journal])) OR ("Journal of strength and conditioning research"[Journal])) OR ("Journal of sports sciences"[Journal])) OR ("Journal of pediatric orthopedics"[Journal])) OR ("Annals of physical and rehabilitation medicine"[Journal])) OR ("Sports health"[Journal])) OR ("Archives of orthopaedic and trauma surgery"[Journal])) OR ("Journal of sport and health science"[Journal]) ) OR ("European journal of applied physiology"[Journal])) OR ("European journal of sport science"[Journal])) OR ("The spine journal : official journal of the North American Spine Society"[Journal])) OR ("International journal of sports medicine"[Journal])) OR ("The Knee"[Journal])) OR ("The Orthopedic clinics of North America"[Journal])) OR ("Physical education and sport pedagogy"[Journal])) OR ("Journal of athletic training"[Journal])) OR ("Calcified tissue international"[Journal]) ) OR ("Sport, education and society"[Journal])) OR ("Journal of orthopaedics and traumatology : official journal of the Italian Society of Orthopaedics and Traumatology"[Journal])) OR ("Journal of orthopaedic trauma"[Journal])) OR ("Journal of orthopaedic research : official publication of the Orthopaedic Research Society"[Journal])) OR ("Journal of biomechanics"[Journal])) OR ("Clinical journal of sport medicine : official journal of the Canadian Academy of Sport Medicine"[Journal])) OR ("EFORT open reviews"[Journal]) ) OR ("Orthopaedics & traumatology, surgery & research : OTSR"[Journal])) OR ("Sports medicine - open"[Journal])) OR ("Clinics in sports medicine"[Journal])) OR ("European physical education review"[Journal])) OR ("The journal of knee surgery"[Journal])) OR ("Injury"[Journal])) OR ("Gait & posture"[Journal])) OR ("Research in sports medicine (Print)"[Journal])) AND ((clinicaltrial[Filter]) AND (2019/12:2020/08[pdat]))

**Data Abstraction**

All reviewers completed training on a minimum of 10 articles to ensure that responses were consistent before starting data abstraction. Data from all included studies wer extracted using preformatted Excel spreadsheets.

**Results**

The search retrieved 175 articles from 27 journals Table S1. All articles were then uploaded into Rayyan (RRID:SCR_017584; 8) for title and abstract screening. Two reviewers (RS, GL) performed title and abstract screening to exclude articles that were obviously not clinical trials, as defined by the ICMJE. The ICMJE defines a clinical trial as any research project that prospectively assigns people or a group of people to an intervention, with or without concurrent comparison or control groups, to study the relationship between a health-related intervention and a health outcome (9). After the title and abstract screening, two independent abstractors (RS, GL, RP) reviewed each full-length, original research article and any available supplemental files. All papers meeting the ICMJE definition of a clinical trial were included. Disagreements were resolved by consensus.

Table S1 Identified top 25% journals that published clinical trials in the time period of interest, the number of identified published articles, and the number of included articles

| **Title** | **Number of articles identified in search** | **Number of included articles** |
| --- | --- | --- |
| Medicine and Science in Sports and Exercise | 22 | 21 |
| Journal of Strength and Conditioning Research | 22 | 21 |
| Bone and Joint Journal | 21 | 18 |
| Journal of Sports Sciences | 13 | 12 |
| British Journal of Sports Medicine | 12 | 12 |
| Knee Surgery, Sports Traumatology, Arthroscopy | 9 | 6 |
| Journal of Bone and Joint Surgery - Series A | 8 | 5 |
| Acta Orthopaedica | 8 | 8 |
| Scandinavian Journal of Medicine and Science in Sports | 8 | 8 |
| American Journal of Sports Medicine | 7 | 7 |
| Journal of Shoulder and Elbow Surgery | 7 | 7 |
| Spine | 6 | 6 |
| Journal of Science and Medicine in Sport | 6 | 6 |
| International Journal of Sports Medicine | 6 | 6 |
| Sports Health | 5 | 5 |
| International Journal of Sports Physiology and Performance | 4 | 4 |
| European Journal of Sport Science | 3 | 3 |
| Journal of Sport and Health Science | 2 | 2 |
| Clinical Orthopaedics and Related Research | 1 | 1 |
| Foot and Ankle International | 1 | 1 |
| Archives of Orthopaedic and Trauma Surgery | 1 | 1 |
| Spine Journal | 1 | 1 |
| Knee | 1 | 1 |
| Journal of Athletic Training | 1 | 1 |
|  | **175** | **163** |

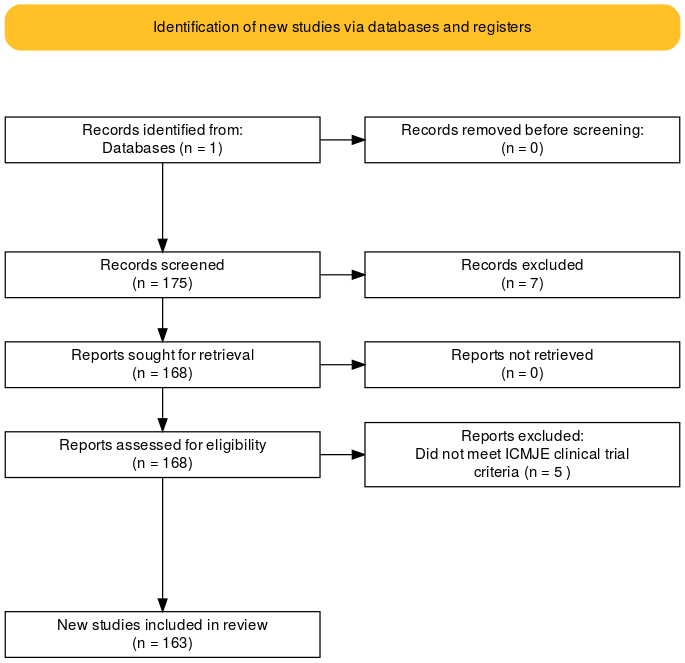

**Figure S1** Flow chart of the study selection process. Seven studies were excluded during the abstract screening because they did not meet the ICMJE clinical trial criteria (n=6) or were the wrong publication type (extended conference abstract; n=1). The flow diagram was created with the ShinyApp for PRISMA 2020 (RRID: 10,11).

References

1.	Daniel J, editor. Sampling essentials: Practical guidelines for making sampling choices. Los Angeles, Calif.: SAGE Publ; 2012. 291 p. eng.

2.	Ghimire S, Kyung E, Kang W, Kim E. Assessment of adherence to the CONSORT statement for quality of reports on randomized controlled trial abstracts from four high-impact general medical journals. Trials. 2012;1377. doi:10.1186/1745-6215-13-77 Cited in: PubMed; PMID 22676267.

3.	Turner L, Shamseer L, Altman DG, Schulz KF, Moher D. Does use of the CONSORT Statement impact the completeness of reporting of randomised controlled trials published in medical journals? A Cochrane review. Syst Rev. 2012;160. doi:10.1186/2046-4053-1-60 Cited in: PubMed; PMID 23194585.

4.	Chess LE, Gagnier J. Risk of bias of randomized controlled trials published in orthopaedic journals. BMC Med Res Methodol. 2013;1376. doi:10.1186/1471-2288-13-76 Cited in: PubMed; PMID 23758875.

5.	Dechartres A, Trinquart L, Atal I, Moher D, Dickersin K, Boutron I, Perrodeau E, Altman DG, Ravaud P. Evolution of poor reporting and inadequate methods over time in 20 920 randomised controlled trials included in Cochrane reviews: research on research study. BMJ. 2017;357j2490. doi:10.1136/bmj.j2490 Cited in: PubMed; PMID 28596181.

6.	Khatkar M, Dhand N. Statulator; 2014.

7.	SCImago (nd). SJR — SCImago Journal & Country Rank [Portal] [Internet]. 2020 [cited 2021 Feb 22]. Available from: https://www.scimagojr.com/journalrank.php?category=2732

8.	Ouzzani M, Hammady H, Fedorowicz Z, Elmagarmid A. Rayyan-a web and mobile app for systematic reviews. Syst Rev. 2016;5(1):210. doi:10.1186/s13643-016-0384-4 Cited in: PubMed; PMID 27919275.

9.	ICMJE. Recommendations for the Conduct, Reporting, Editing and Publication of Scholarly Work in Medical Journals [Internet]. 2020 [cited 2020 Aug 4]. Available from: http://www.ICMJE.org.

10.	Haddaway NR, McGuinness L. PRISMA2020: R package and ShinyApp for producing PRISMA 2020 compliant flow diagrams: Zenodo; 2020.

11.	Page MJ, McKenzie JE, Bossuyt PM, Boutron I, Hoffmann TC, Mulrow CD, Shamseer L, Tetzlaff JM, Akl EA, Brennan SE, Chou R, Glanville J, Grimshaw JM, Hróbjartsson A, Lalu MM, Li T, Loder EW, Mayo-Wilson E, McDonald S, McGuinness LA, Stewart LA, Thomas J, Tricco AC, Welch VA, Whiting P, Moher D. The PRISMA 2020 statement: an updated guideline for reporting systematic reviews. BMJ. 2021;372n71. doi:10.1136/bmj.n71 Cited in: PubMed; PMID 33782057.
